# A structured model for COVID-19 spread: modelling age and healthcare inequities

**DOI:** 10.1101/2020.05.17.20104976

**Authors:** A. James, M. J. Plank, R. N. Binny, A. Lustig, K. Hannah, S. C. Hendy, N. Steyn

## Abstract

We use a stochastic branching process model, structured by age and level of healthcare access, to look at the heterogeneous spread of COVID-19 within a population. We examine the effect of control scenarios targeted at particular groups, such as school closures or social distancing by older people. Although we currently lack detailed empirical data about contact and infection rates between age groups and groups with different levels of healthcare access within New Zealand, these scenarios illustrate how such evidence could be used to inform specific interventions. We find that an increase in the transmission rates amongst children from reopening schools is unlikely to significantly increase the number of cases, unless this is accompanied by a change in adult behaviour. We also find that there is a risk of undetected outbreaks occurring in communities that have low access to healthcare and that are socially isolated from more privileged communities. The greater the degree of inequity and extent of social segregation, the longer it will take before any outbreaks are detected. Well-established evidence for health inequities, particularly in accessing primary healthcare and testing, indicates that Māori and Pacific peoples are at higher risk of undetected outbreaks in Aotearoa New Zealand. This highlights the importance of ensuring that community needs for access to healthcare, including early proactive testing, rapid contact tracing, and the ability to isolate, are being met equitably. Finally, these scenarios illustrate how information concerning contact and infection rates across different demographic groups may be useful in informing specific policy interventions.

## 1. Introduction

The COVID-19 outbreak originated in Wuhan China in late 2019 (World Health Organisation, 2020a) before spreading globally to become a pandemic in March 2020 (World Health Organisation, 2020b). The human population currently lacks immunity to COVID-19, a viral zoonotic disease with reported fatality rates that are of the order of 1% (Verity *et al*., 2020). Many countries have experienced community transmission after undetected introductions of the disease by travellers exposed in China. This has led to exponential growth of new infections in many countries, even as China, through the use of strong controls and rapid testing and tracing, has managed to control the disease.

The effects of the COVID-19 pandemic do not impact people or communities equally. For instance, there is strong evidence internationally that the severity of symptoms and the infection fatality rates (IFR) vary by orders of magnitude across age groups, with estimates of the IFR in children (aged 0-9) as low as 0.0016% compared to 7.8% in adults over 80 years of age (Verity *et al*., 2020). Furthermore, overcrowded living conditions (House & Keeling, 2009) and employment in high-contact service occupations (Koh, 2020) are also likely to be risk factors for infection, potentially increasing the burden of COVID-19 for socio-economically disadvantaged communities.

Indeed, in Singapore, structural socio-economic and healthcare inequities have fuelled a secondary COVID-19 outbreak. Despite its government’s early claims to have contained the spread of COVID-19, a large secondary outbreak emerged in Singapore’s migrant worker community in April (Singapore Ministry of Health, 2020). These communities are housed in crowded dormitory precincts where self-isolation is difficult and report high levels of job insecurity, which creates structural disincentives to reporting illness and to enact self-isolation (HOME, 2020). The experiences of migrant workers in Singapore, which include overcrowded housing, and lack of access to primary healthcare or sick leave, are an indicator of the importance of equitable epidemic control measures that account for the lived experiences of all.

In Aotearoa New Zealand, inequities in health and healthcare (Waitangi Tribunal, 2019) have been shown to substantially increase relative fatality rates for Māori and Pacific (Steyn *et al*., 2020). Furthermore, Māori and Pacific people are more likely to live in crowded conditions than European New Zealanders (Stats NZ, 2018). Population count data also suggests that socio-economic status in Aotearoa New Zealand is a key indicator for a community’s ability to reduce contacts during working hours, with wealthier communities able to work more easily from home (Data Ventures, 2020). This combination of structural inequities has the potential to lead to both higher transmissibility and higher infection fatality rates for COVID-19, resulting in higher risks for Māori and Pacific populations in New Zealand.

In this paper we introduce a structured branching process model similar to that of Davies *et al*. (2020a) that allows for heterogeneous contact networks among different age groups and between communities in the presence of healthcare inequities. Each group has its own parameter set to describe the average reproduction number, the probability of being asymptomatic, and the level of access to testing for individuals within the group. The connections between such groups are described by a contact matrix, which determines the relative likelihood of an infected individual from group *i* infecting someone from group *j*.

The main purpose of this study is not to produce detailed forecasts of COVID-19 cases in different age or other demographic groups. Although estimates of contact rates between groups corresponding to 5-year age bands exist for New Zealand (Prem *et al*., 2020), these do not account for the effects of behavioural changes, social distancing and other interventions in response to COVID-19. In addition, New Zealand has had a relatively small number of COVID-19 cases (1), which mean it is difficult to estimate contact rates across finely stratified age groups. Stratifying the model at this level would therefore introduce too many undetermined parameters.

Instead, we aim to develop a coarse-grained model that can compare control interventions that differentially affect different broad age groups (e.g. school closures) and can help identify potential risks to particular communities. We estimate contact rates between broad age classes qualitatively from available data and test the sensitivity of the model to changes in the structure of the contact rates. The model is formulated in a mathematically general way that allows it to be applied to any number of population groups. This will be useful when combined with robust New Zealand-specific data on contact rates between fine-grained groups at varying Alert Levels and epidemiological properties of COVID-19 within these groups. In this work, we use this model to explore two scenarios: the first is an age-structured model that splits the population into three groups based on age, which we use to investigate the effects of school closures, while the second splits the population into two communities in the presence of healthcare inequities.

## 2. Methods

We use a branching process simulated in 1 day time steps to model the number of infections in each group, with initial seed cases representing overseas arrivals. The number and timing of these seed cases was chosen to replicate real case data (see section 2.1).

For each scenario we segment the population into *N*_*G*_ mutually exclusive groups. Within each group, individuals are assumed be homogeneous, with the exception of their individual reproduction number, which is heterogeneous and randomly distributed with a group-specific mean (see below).

Infected individuals fall into two categories: (i) those who show clinical symptoms at some point during their infection; and (ii) those who are asymptomatic or subclincal for the duration of their infection. Each new infection is randomly assigned as subclinical with probability 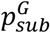 and clinical with probability 1 − 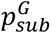, independent of who infected them. This categorisation applies for the full duration of the infectious period, i.e. the clinical category includes pre-symptomatic individuals who later go on to develop clinical symptoms. The probability of being subclinical 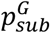 may vary between groups.

Clinical individuals have an initial period during which they are either asymptomatic or have sufficiently mild symptoms that they have not isolated. During this period, their infectiousness is as shown by the blue curve in Figure 1. Once they develop more serious symptoms, they have a probability of being detected 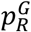, and thus are isolated. In this case their infectiousness reduces to *c*_*iso*_ = 65% (Davies *et al*., 2020a) of the value it would have without isolation (green curve in Figure 1). This represents a control policy of requiring symptomatic individuals to self-isolate. Subclinical individuals are never isolated.

**Fig 1.**
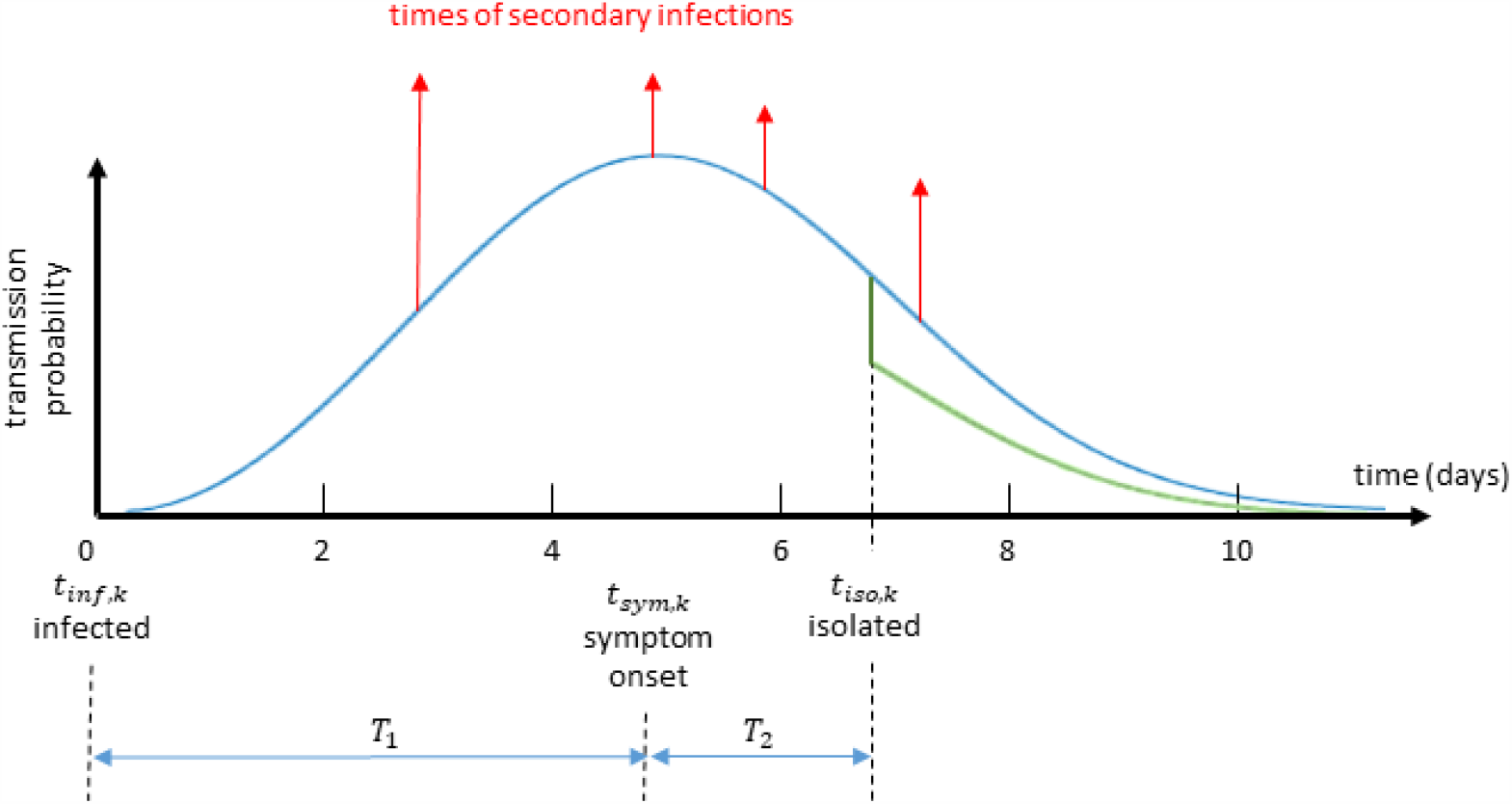
Schematic diagram showing the timeline and transmission probability of a typical case. In the early stages secondary infections are unlikely. Red arrows show the exposure times of new secondary infections. After isolation the transmission probability is reduced by a fixed proportion (green curve). Subclinical infections are not isolated and follow the shape of the blue curve throughout, but with a lower overall infectiousness. Time from infection to isolation is the sum of two random variables *T*_1_ and *T*_2_ drawn from the distributions shown in Table 1.

The average reproduction number 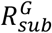 of subclinical individuals in any group *G* was assumed to be 50% of the average reproduction number 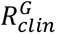 of clinical individuals that group. This reflects lower infectiousness of subclincal cases (Davies *et al*., 2020a). Individual heterogeneity in transmission rates was included by setting 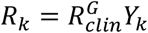 for clinical individuals in group *G* and 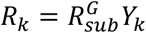 for subclinical individuals in group *G*, where *Y*_*k*_ is a gamma distributed random variable with mean 1 and variance 2 (Lloyd-Smith *et al*., 2005). In the absence of case isolation measures (see section 2.2), each infected individual *k* causes a randomly generated number *N*_*k*_∼*Poisson*(*R*_*k*_) of new infections.

**Table 1.** The parameters used in the model and their source. See section 2.1 for a description of how parameters were estimated from New Zealand case data.

| Parameter | Value | Source |
| --- | --- | --- |
| Distribution of generation times | $Weibull(shape = 2.83, scale = 5.67)$ | Feretti et al. (2020) |
| Distribution of exposure to onset (days) | $T_1 \sim \Gamma(shape = 5.8, scale = 0.95)$ | Lauer et al. (2020) |
| Distribution of onset to isolation (days) (from data) | $T_2 \sim Exp(2.18)$ | Estimated from data |
| Distribution from isolation to reporting | $T_3 \sim \Gamma(shape = 0.95, scale = 3.5)$ | Estimated from data |
| Relative infectiousness of subclinical cases | $R_{sub}^G / R_{clin}^G = 50\%$ | Davies et al. (2020a) |
| Relative infectiousness after isolation | $c_{iso} = 65\%$ | Davies et al. (2020a) |
| Parameters for age-structured scenario |  |  |
| Reproduction number for clinical infections (no case isolation or control) | $R_{clin} = 4.6, 3, 1.3$ | Estimated from age-adjusted contact rates (Li et al., 2017) |
| Proportion of subclinical infections | $p_{sub} = 0.8, 0.33, 0.2$ | Davies et al., 2020b |
| Proportion of cases detected | $p_R = 0.75, 0.75, 0.75$ | Price et al. 2020 |
| Contact probabilities between groups at control levels 1-2 | $\Lambda = \begin{matrix} 0.6 & 0.325 & 0.075 \\ 0.1 & 0.825 & 0.075 \\ 0.1 & 0.325 & 0.575 \end{matrix}$ | See text |
| Contact probabilities between groups at control levels 3-4 | $\Lambda = \begin{matrix} 0.475 & 0.475 & 0.05 \\ 0.3 & 0.65 & 0.05 \\ 0.05 & 0.3 & 0.65 \end{matrix}$ | |
| Contact probabilities between groups at control levels 3-4 (schools open) | $\Lambda = \begin{matrix} 0.6 & 0.325 & 0.075 \\ 0.3 & 0.65 & 0.05 \\ 0.05 & 0.3 & 0.65 \end{matrix}$ | |
| Contact probabilities between groups at control levels 3-4 (schools open and increased parental interaction) | $\Lambda = \begin{matrix} 0.6 & 0.325 & 0.075 \\ 0.1 & 0.825 & 0.075 \\ 0.05 & 0.3 & 0.65 \end{matrix}$ | |
| Transmission rates at levels 1-4 | $C = 1, 0.7, 0.45, 0.2$ | See text |
| Parameters for scenario structured by access to healthcare |  |  |
| Reproduction number for clinical infections (no case isolation or control) | $R_{clin} = 3, 3$ | Davies et al. (2020a) |
| Proportion of subclinical infections | $p_{sub} = 0.33, 0.33$ | Davies et al. (2020a) |
| Proportion of cases detected | $p_R = 0.75, 0.05$ | |
| Contact probabilities between groups at control levels 1-4 | $\Lambda = \begin{matrix} 0.99 & 0.01 \\ 0.01 & 0.99 \end{matrix}$ | See text |
| Transmission rates at levels 1-4, good health care access group | $C = 1, 0.7, 0.3, 0.2$ | |
| Transmission rates at levels 1-4, lower health care access group | $C = 1, 0.7, 0.6, 0.4$ | |

The time between an individual becoming infected and infecting another individual, the generation time *T*_*G*_, is Weibull distributed with mean 5.0 days and standard deviation of 1.9 days (Ferretti *et al*., 2020). The infection times of all *N*_*i*_ secondary infections from individual *i* are independent, identically distributed random variables from this distribution (see Figure 1). An alternative log normal distribution and a Gamma distribution were investigated (Nishiura *et al*., 2020). Whilst they each required small changes in control levels to achieve a good fit with case data, they did not change the results significantly. For computational convenience, all individuals are assumed to be no longer infectious 30 days after being infected. In practice, individuals have very low infectiousness after about 12 days because of the shape of the generation time distribution (Fig. 1).

The time between infection and isolation the sum of two random variables *T*_1_ and *T*_2_. The random variable *T*_1_ represents the time from infection to onset of symptoms and has a gamma distribution with mean 5.5 days and shape parameter 5.8 (Lauer *et al*., 2020). The random variable *T*_2_ is the time from symptom onset to isolation and is assumed to be exponentially distributed. There is an additional delay *T*_3_ from isolation to reporting, which is modelled as a gamma distribution. Parameters for the distributions of *T*_2_ and *T*_3_ were estimated from New Zealand case data (see section 2.1). The model does not explicitly include a latent period, however, the shape of the Weibull generation time distribution (Figure 1) captures this effect, giving a low probability of a short generation time between infections.

The model is simulated using a time step of *δt* = 1 day. In the time step [*t, t* + *δt*], infectious individual *k* produces a Poisson distributed number of secondary infections with mean

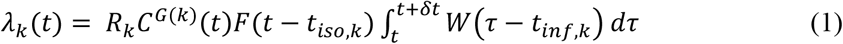

where *R*_*k*_ is the mean number of secondary infections from individual *k, W* is the probability density function of the Weibull distributed generation time (see Table 1), *t*_*inf,k*_ and *t*_*iso,k*_are the infection time and isolation time respectively for individual *k, C*^*G*(*k*)^(*t*) is the control effectivity at time *t* (see section 2.2) for the group *G*(*k*) to which individual *k* belongs, and *F*(*t*) is a function describing the reduction in infectiousness due to isolation:

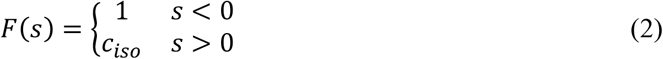

A contact matrix Λ gives the probability Λ_*GH*_ that a secondary infection originating from group *G* will be in group *H*, with ∑_*H*_ Λ_*GH*_ = 1. New cases infected by an individual in group *G* are distributed across groups according to these probabilities. All individuals infected during time step [*t, t* + *δt*] are assigned infection time *t*_*inf,k*_ = *t* + *δt*.

### 2.1 Case data

Model simulations were seeded with the *N*_*int*_ = 501 New Zealand cases that had a history of international travel and a known international arrival date prior to 10 April 2020. After 10 April 2020, all international arrivals to New Zealand were required to spend 14 days in government-managed quarantine (Jefferies et al., 2020). For the *N*_*int*_ international seed cases, the infection date was estimated backwards from the date of onset of symptoms (distribution shown in Table 1). For cases that did not include an onset date, the infection date was backdated from the arrival date. Cases missing an isolation date were assumed to remain fully infectious for the whole infectious period. Secondary infections that occurred before arrival in New Zealand were ignored (i.e. the right-hand side of Eq. (1) was set to zero for values of *t* preceding date of arrival). Cases that were flagged as having an international travel history but missing an arrival date were assumed to have arrived on the same date at becoming infected, so all their secondary infections were included. To allow for the fact that the case data only includes clinical cases, an additional number *N*_*int,sub*_∼*Poisson*(*N*_*int*_*p*_*sub*_/(1 − *p*_*sub*_)) of subclinical seed cases were added. The arrival, onset and isolation dates for these subclinical seed cases were approximated by random sampling with replacement from the clinical seed cases.

The times from symptom onset to isolation and from isolation to reporting were calculated for the *N*_*dom*_ = 713 domestically acquired cases of COVID-19 reported in New Zealand up to 7 April 2020 (see Table 1). The mean of the distribution of *T*_2_ was set to be the mean of the observed times from symptom onset to isolation. The parameters of the gamma distribution for *T*_3_ were set to be the maximum likelihood estimates using the observed times from isolation to reporting. The international cases were assigned the actual reporting date as recorded in the data.

### 2.2 Control sub-model

Population-wide control interventions were modelled via the function *C*^*G*^(*t*), which represents the transmission rate for infected individuals in group *G* relative to no population-wide control. Population-wide control interventions include closure of schools, universities or non-essential businesses, restrictions on large gatherings and domestic travel, social distancing measures and stay-at-home orders. New Zealand’s control measures are based on a scale of Alert Levels ranging from 1 to 4. Level 1 controls are largely focussed on border measures, while Level 4 is a strong lockdown of most activities except for ‘essential services’. Alert Level 2 was announced on 21 March and this was raised to Alert Level 3 on 23 March and Alert Level 4 on 25 March. We do not attempt to model the short periods of time at Alert Levels 2 and 3; instead we assume that Level 2 was in place up to 25 March, with an effective reproduction number consistent with the data up to this point. After 25 March, transmission rates are reduced to represent Alert Level 4, which continues until the end of 27 April.

Reductions in the transmission rates at Alert Levels are based on modelling literature (Moss *et al*., 2020; Kissler *et al*., 2020; Jarvis *et al*., 2020) and empirical estimates of the effective reproduction number in international data (Flaxman *et al*., 2020; Hsiang *et al*., 2020; Binny *et al*., 2020) and provide a reasonable fit with the New Zealand data. Estimates for Level 3 transmission reduction should be treated with caution at this stage and estimates used here may not be representative. The control level *C*^*G*^ can also be varied across the different population groups. This can capture a range of effects specific to a particular group, for example the practicalities of implementing social distancing in a school or early childhood setting, the difficulties in isolating older people in multi-generational households or maintaining an isolated household bubble in overcrowded housing.

### 2.3 Contact sub-model and effective reproductive number

For individuals in group *G*, the average reproduction number without case isolation is

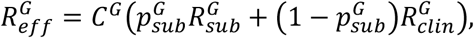

where *C*^*G*^ ≤ 1 is the transmission rate in group *G* relative to transmission with no control measures. The average number of secondary infections in group *H* caused by a randomly chosen index case in group *G* is 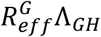. The overall population effective reproduction number *R*_*eff*_ is the dominant eigenvalue of the 2*N*_*G*_ × 2*N*_*G*_ block matrix

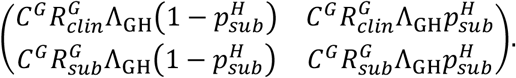

where each of the four blocks is a *N*_*G*_ × *N*_*G*_ matrix over all combinations of *G* and *H*. The contact probability matrix Λ is chosen to model particular scenarios to compare the relative efficacy of different control interventions or to help identify potential high-risk groups. This is described in detail for each scenario below and the associated values chosen for Λ are shown in Table 1.

### 2.4. Sensitivity analysis

We have made a number of simplifying assumptions in developing and parameterising the model. We have tested the sensitivity of model results to variations in some of the model parameters whose values are uncertain or context-dependent. These include the proportion and relative infectiousness of subclinical cases, the heterogeneity in individual reproduction numbers, the mean generation time, and the values in the contact matrix. Any change in parameters that results in a change in the overall population reproduction number *R*_*eff*_ causes a corresponding change in the trajectory of case numbers. However, if the value of *R*_*eff*_ is fixed, the model is robust to these parameters and the qualitative results described in the scenarios above are not affected. Increasing heterogeneity in individual reproduction numbers increases variation between independent stochastic realisations and increases the probability of ultimate extinction (Lloyd-Smith *et al*., 2005). This is important in scenarios that consider elimination of the virus as a potential outcome, but this was not the primary focus of our work here. Increasing mean generation times slows the spread of the virus and, if the model were then calibrated against empirical case data, this would require a larger value of *R*_0_ (and by implication greater relative reduction in *R*_0_ at each Alert Level). However, the distinction between short generation time and low *R*_0_ versus longer generation time and higher *R*_0_ is not crucial in scenarios where there is no significant herd immunity. Model results are insensitive to moderate changes in the contact matrix but will change slightly if very extreme values are used.

## 3. Scenarios

### 3.1 Age-structured scenario

In these scenarios, we consider the effect of school closure on disease spread. Detailed data about the effects of lockdown and other COVID-19 response measures on age-structured contact rates are not yet reliably available. Although this may partially emerge from a combination of data sources including employment data, telecommunications data, calibrating this with New Zealand household size data and case and contact tracing data is beyond the scope of this paper. In the absence of such data, we have made a number of assumptions including the rates of transmission between under 19 year olds and adults and with groups of under 19 year olds. Nonetheless, the opening and closing of schools is an important policy choice, so we want to explore the possibility of answering this question using an age-structured model.

As such, we consider a population split into three age groups: under 19 year olds, 19 – 65 year olds and over 65s. Data shows that these three age groups have very different rates of clinical severity to COVID-19 (Verity *et al*., 2020) and we assume that the probability of being subclinical is 80%, 33% and 20% respectively based on estimates by Davies *et al*. (2020b). We assume that all age groups have the same level of healthcare access and clinical cases are detected equally in all groups. Seed cases are assigned to an age group according to the NZ data.

At Alert Levels 1 and 2, we assume that 50% of contacts between groups occur according to the relative size of each group in the population (here under 19s - 20%, 19-65 year olds – 65% and over 65s – 15%). The remaining 50% of contacts are within group. This is similar to the age-structured contact model of Prem *et al*. (2020) although with a coarser age stratification. It is also consistent with projections for age-stratified contact rates for New Zealand (Prem *et al*., 2017) aggregated into coarser age groups. We assume that older people have fewer contacts with other groups: only 5% of secondary cases originating from children are in older people, with remaining secondary cases from children split evenly between other children and adults. This models an average child’s isolation bubble as containing a similar number of adults and other children. The majority (65%) of secondary infections originating from adults are still in other adults, with only 5% in older people and the remainder (30%) with children. Finally, secondary cases originating from older people occur predominantly (65%) in other older people, only 5% in children and the remainder (30%) in adults.

The mean reproduction number, in the absence of control measures, for clinical cases, 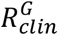, is assumed to be 3 (Davies *et al*., 2020a). For the children and older people groups, it is scaled according to the expected number of face-to-face contacts given in the 2017 International Social Survey Programme for New Zealand (Li *et al*., 2018). This results in clinical cases among children having a much higher average reproduction number and among older people a much lower one. We assume that control measures at Alert Levels 3 and 4 produce same relative in transmission from all three age groups.

This model fits well with data on the number of cases in NZ in each age group over time (see Figure 2, red lines). The model results are most dependent on the proportion of subclinical and undetected cases in each group (as described above, these were sourced from literature not tuned to fit the model to data). Notwithstanding extreme choices, the results are relatively insensitive to changes in the contact matrices. This is in part due to the large number of international seed cases in the model but also suggests that age-specific epidemiological parameters play a more important role than the details of contact rates between age groups. This suggests that the results seen here might remain robust once further data is available to inform actual contact and/or transmission rates.

**Fig 2.**
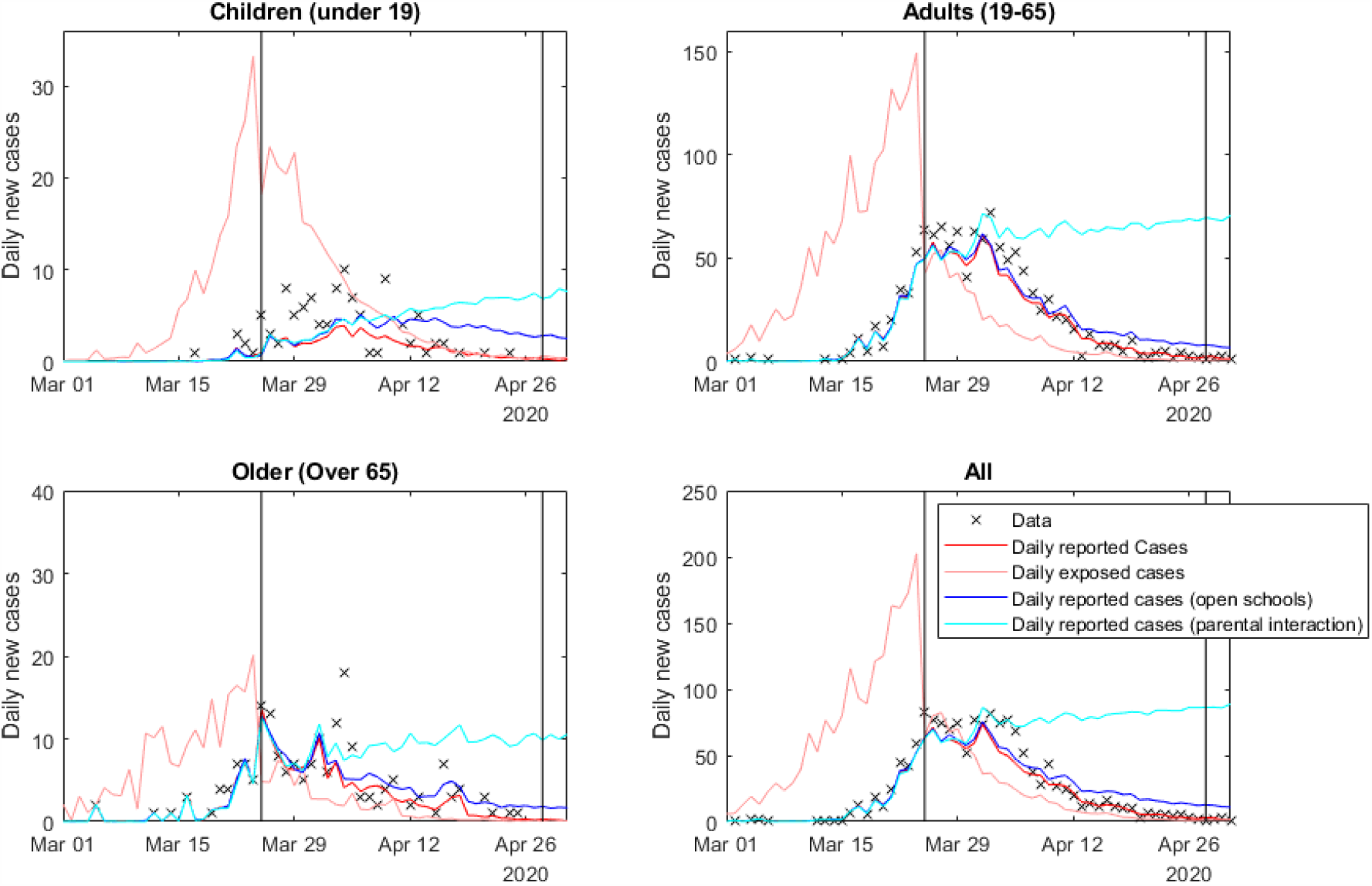
Results of the age-structured model. Red lines show the daily number of reported cases; pink lines show the number of newly infected cases each day. These reflect the actual situation where schools were closed from March 25. The blue line shows the reported daily cases if schools had remained open, while the light blue line shows the reported cases if school openings increased parental contacts. Results are averaged over 500 independent, identically initialised realisations of the stochastic process.

We now consider hypothetical school closures as a test of the model. Different countries have implemented very different policies for schools. As very little is known about overall transmission rates at Alert Level 3, we consider a counterfactual scenario in which schools remained open during Alert Level 4. The blue line in Figure 2 shows the model output if the average reproduction number for the under 19s group during Levels 4 was at the much higher level associated with Level 3. We also keep the first row of the contact matrix the same as at Levels 1 and 2 (see Table 2). This means that a higher proportion of secondary infections originating from children are in other children. We see that relaxation of control for children has only a small effect on the total number of cases and the overall population effective reproduction number is still less than 1 (Table 1). However, the light blue line shows the effect of opening schools with an associated increase in adult, i.e. parental, interaction. Here, the average reproduction number for adults is also set to the higher level under Alert level 3 (Table 1) and the adult row of the contact matrix reverts back to the Level 1-2 values (Table 1). This has a much greater effect on the overall case numbers than the increased transmission rates of children alone. The overall population effective reproduction number is above 1 in this scenario (Table 2).

**Table 2.** Summary of transmission coefficients (i.e. control level) by group and effective reproduction numbers in the two scenarios.

| | $R_{eff}$ and associated reduction in transmission rates from Level 1 | | | | |
| --- | --- | --- | --- | --- | --- |
|  | Level 2 | Level 3 | Level 4 | Level 4<br>(schools open) | Level 4<br>(schools open,<br>parental interaction) |
| Age-structured model |  |  |  |  |  |
| Children | 1.97 (70%) | 1.27 (45%) | 0.56 (20%) | 1.27 (45%) | 1.27 (45%) |
| Adults | 1.75 (70%) | 1.13 (45%) | 0.5 (20%) | 0.5 (20%) | 1.13 (45%) |
| Elderly | 0.83 (70%) | 0.53 (45%) | 0.24 (20%) | 0.24 (20%) | 0.24 (20%) |
| $R_{eff}$ | 1.66 | 1.07 | 0.47 | 0.86 | 1.06 |
| Model structured by access to healthcare |  |  |  |  |  |
| High detection<br>($p_R = 0.75$) | 1.75 (70%) | 0.75 (30%) | 0.5 (20%) | -- | -- |
| Low detection<br>($p_R = 0.05$) | 1.75 (70%) | 1.5 (60%) | 1 (40%) | -- | -- |
| $R_{eff}$ | 1.75 | 1.49 | 0.99 | -- | -- |

### 3.2 Scenario structured by inequity in access to healthcare

Singapore has seen a second outbreak of infections after appearing to contain an initial outbreak (Singapore Ministry of Health, 2020). This second wave emerged in a migrant worker community with low socioeconomic indicators, overcrowded housing, and less access to healthcare and testing. The community also faces strong economic disincentives to seek healthcare (HOME, 2020). In these circumstances, the outbreak has been difficult to contain and was likely detected late. In this section we want to explore scenarios that capture aspects of this situation.

We do not attempt to model the multitude of relevant socioeconomic and demographic variables in detail, but instead explore the effect of different transmission and case detection rates in a simplified scenario that captures certain aspects of these. We split the population into two groups, each with a different level of access to healthcare and COVID-19 testing and diagnosis. To simplify the model, we assume each group has the same proportion of subclinical cases, although we note that COVID-19 severity and access to healthcare are likely to have common covariates for example derivation index and ethnicity (Steyn 2020). We assume that in the group with good access to healthcare, 75% of clinical cases are detected. Individuals in this group are less likely to be in overcrowded housing and more likely to have jobs that are compatible with social distancing or working from home. For these reasons, we assume that transmission rates in this group drop significantly under Levels 3 and 4 control.

We assume that the group with poorer access to healthcare is smaller in size and has a much lower clinical case detection rate of 5%. This should be interpreted as meaning that only the most clinically severe cases are detected (although variation in severity of clinical cases in not explicitly modelled). We also investigate the scenario where transmission rates in this group do not decrease as much as in the first group during Alert Levels 3-4 due to, for example, overcrowded housing and job security issues precluding self-isolation during illness. These parameter choices represent a simple model of inequities in the healthcare system and socioeconomic correlates, rather than actual estimates of case detection and transmission rates for particular groups. The motivation for this is to identify potential factors and regions of parameter space where large outbreaks could go undetected for a long period, for example as occurred in Singapore.

For simplicity, all seed cases are in the larger group with high levels of healthcare access. In the first scenario, we assume that both groups have the same reduction in transmission rates at any given Alert Levels (Figure 3, red lines). This models a desired outcome of a policy intervention, whereby an effective control strategy takes into account the differing contexts for different sectors of society in order to achieve equitable outcomes. In this scenario, the epidemic is successfully controlled. In the second scenario (Figure 3, blue lines), we assume that control measures are less effective in the group with poor healthcare access. Throughout the Level 4 period, reported case numbers are almost identical but, due to low access to testing, there is a rapidly growing number of undiagnosed infections in the group with poor healthcare access. As a result, once overall control is relaxed at the end of Level 4, case numbers start to rise rapidly in both groups.

**Fig 3.**
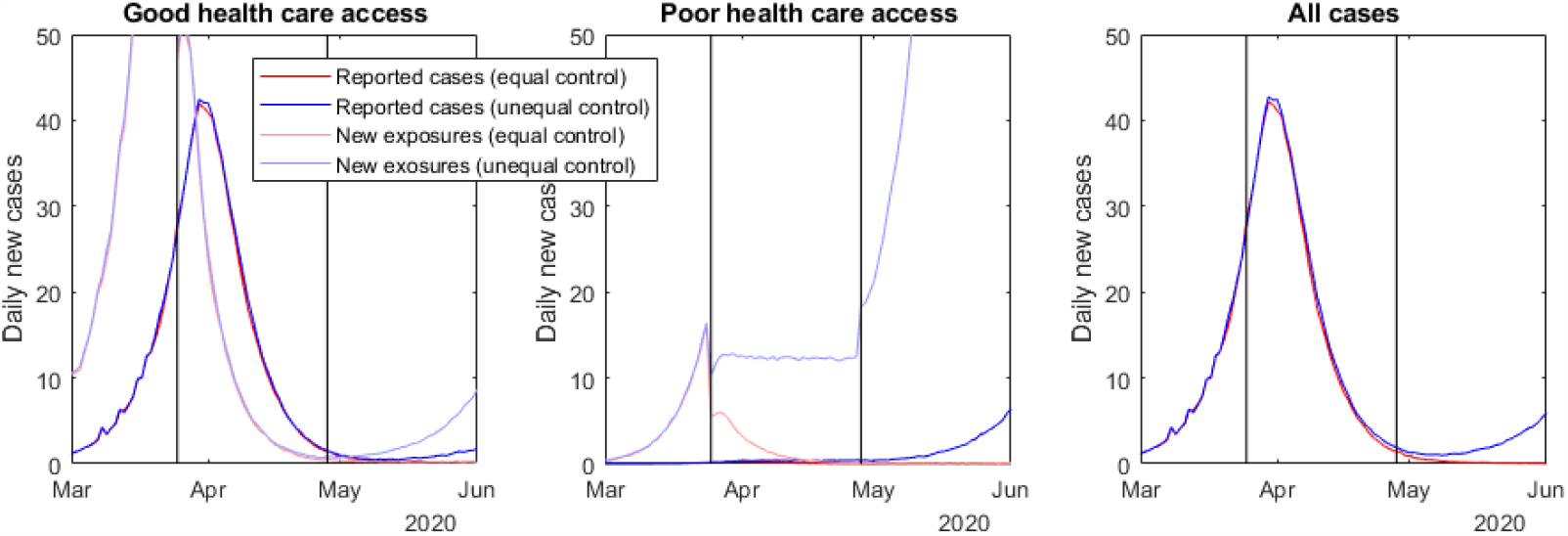
The results of a model structured by access to healthcare, where cases are less likely to be detected in one group because of poor access to testing and diagnosis. The red line illustrates a scenario where control measures are equally effective for both groups. The blue line shows a scenario where control measures do not reduce transmission rates for the group with poor access to healthcare, eventually leading to an outbreak in both populations. Pale lines show the corresponding daily number of new exposures. Results are averaged over 500 independent, identically initialised realisations of the stochastic process.

One of the key risk factors driving this result is the low contact rate between the two groups. If this contact rate is large, a growth in the number of cases in one group will quickly lead to cases in the other group and outbreaks will be promptly detected through the higher testing rate of the group with better healthcare access. This suggests that the highest-risk groups are those that have both low levels of access to healthcare and limited contact rates with other groups. Although Auckland New Zealand can be considered a diverse city in aggregate, there is segregation by ethnicity, particularly for Pacific peoples (Manley *et al*., 2015; Salesa, 2017). Pacific peoples in Auckland also experience higher rates of overcrowding (Schluter *et al*., 2007) and high rates of unmet healthcare needs (Ryan *et al*., 2019). This study suggests that Pacific communities in Auckland are at risk of large secondary outbreaks if structural inequities in healthcare are not addressed.

## 4. Discussion

We have introduced a stochastic model of COVID-19 that can account for age structure and inequities in healthcare access. Although we lack detailed data on contact and transmission rates between some groups at varying Alert Levels, we have made a number of assumptions that allow us to explore the potential effects of omitting these in the original model. In particular, we have looked at two sets of scenarios where age and healthcare access are likely to be important and which would not be well described by a homogeneous population model. Firstly, we considered school closures, where it is important to capture differences in infection and contact rates in adults and school pupils. Secondly, we looked at a scenario where socio-economic and healthcare inequities lead to a large secondary outbreak in a high-risk community, much like that which occurred in migrant worker communities in Singapore. The effective reproduction numbers *R*_*eff*_ for these scenarios are shown in Table 2. The table shows that specific interventions can have a significant effect on the effectiveness of control under Alert Levels 3 and 4.

There are some important caveats to this work. While we have endeavoured to consider scenarios that are consistent with existing evidence, for the most part the current evidence base is currently too thin to draw firm conclusions, particularly around school closures. Nonetheless, it is anticipated that better information on contact rates for various groups in New Zealand will be available soon. This study illustrates how such information will allow us to better understand the risk factors for specific communities in New Zealand and how specific control measures might reduce the impact on these communities.

The age-structured model used very coarse age classes. This allows investigation of school closures or other policy interventions that differentially affect transmission in different age groups. However, these coarse age classes are likely to contain significant heterogeneity in variables such as occupation, as well as masking finer-scale contact structures. The model framework allows finer age classes once sufficient data is available to parameterise contact rates under different control interventions and epidemiological parameters (e.g. proportion of subclinical cases) for these groups. We examined age structure and inequities in access to healthcare as two separate scenarios. These could be combined by stratifying the population by age and level of healthcare access simultaneously. Again this would require more detailed data to parameterise the contact structure for the increased number of groups this will require.

Nonetheless, structuring the model by levels of inequity in healthcare, which itself may be a consequence of differing levels of socioeconomic deprivation and/or racial discrimination, is important in guiding New Zealand’s COVID-19 response. The New Zealand government has an obligation under Te Tiriti o Waitangi to deliver equitable health outcomes for Māori and other population groups (Waitangi Tribunal 2019). Māori and Pacific peoples and communities experience inequities in health and healthcare access that increase the risks of infection with COVID-19 and magnify the impacts of the disease (McLeod *et al*., 2020). For example, Māori and Pacific peoples are disproportionately affected by crowded housing (Schluter *et al*., 2007; StatsNZ, 2018), have shorter life expectancy (StatsNZ, Infoshare), higher prevalence of comorbid health conditions (Chan *et al*., 2020; Coppell *et al*., 2013; Telfar-Barnard & Zhang 2018; Ministry of Health, 2019a), and are expected to suffer a higher infection fatality ratio (Steyn *et al*., 2020). Particularly relevant to this study, Māori and Pacific peoples have poorer access to healthcare services and higher unmet health needs (Ministry of Health, 2019a, 2019b). Our model results show that communities with poor access to healthcare and low contact rates with other population groups are at high risk of suffering a large outbreak, which could remain undetected for a long time. This could include urban Pacific communities, as well as remote communities, including rural Māori communities. The government has a responsibility to work with these communities to ensure that their healthcare needs are being met by its COVID-19 response.

There is a growing body of the international evidence that socioeconomic disparities do indeed magnify the impacts of the COVID-19. In the United States, for instance, predominantly African American communities have faced infection rates three times those of European American communities and death rates six times as high (Yancy, 2020). A report from the United Kingdom suggests that ethnic minority groups are at significantly higher risk from COVID-19 (ICNARC, 2020). We have not modelled potential differences in clinical severity (e.g. hospitalisation rates) or transmission rates across groups with different levels of access to healthcare. This will be important to do, especially given these variables are likely to be correlated (Steyn *et al*., 2020), amplifying the impact of COVID-19 in high-risk groups. This is therefore a crucial future objective to inform healthcare planning, prioritisation of high-risk groups and addressing inequities in the healthcare system.

## Data Availability

No new data is referred to in the manuscript.

## Acknowledgements

The authors are grateful to Samik Datta, Nigel French, Markus Luczak-Roesch, Melissa McLeod, Anja Mizdrak, Fraser Morgan, Matt Parry and one anonymous reviewer for comments on an earlier version of this manuscript, and to Dion O’Neale and Emily Harvey for sharing data on age-specific contact rates in New Zealand. The authors also thank the Ministry of Health’s Technical Advisory Group Epidemiology Subcommittee for their comments and suggestions. The authors acknowledge the support of StatsNZ, ESR, and the Ministry of Health in supplying data in support of this work and financial support from Te Pūnaha Matatini.

